# Disrupted white matter integrity in treatment-resistant schizophrenia

**DOI:** 10.1101/19010777

**Authors:** Carolyn B. McNabb, Meghan E. McIlwain, Valerie M. Anderson, Robert R. Kydd, Frederick Sundram, Bruce R. Russell

## Abstract

Treatment response in schizophrenia is heterogeneous and has been posited to divide into three distinct subcategories: treatment-responsive (first-line responders; FLR), treatment-resistant (TRS, responding to clozapine), and ultra-treatment-resistant schizophrenia (UTRS, requiring augmented antipsychotic therapy). Previous work suggests that white matter abnormalities drive antipsychotic resistance but little work has been carried out to identify differences between those with TRS and those with UTRS. The current study aimed to establish whether differences in white matter structure are present across both treatment-resistant subtypes of schizophrenia or if UTRS is distinct from TRS. Diffusion-weighted images were acquired for 18 individuals with TRS, 14 with UTRS, 18 FLR and 20 healthy controls. Measures of fractional anisotropy (FA), mean diffusivity (MD), radial diffusivity (RD) and parallel diffusivity (PD) were obtained using tract-based spatial statistics. Analysis of variance (ANOVA) and post-hoc between-groups t-tests interrogating differences were conducted for each white matter measure. Those with TRS had lower FA than healthy controls across widespread regions of the brain, including the superior longitudinal fasciculus, corpus callosum, thalamic radiation, corticospinal tract, internal capsule, corona radiata and fronto-occipital fasciculus (p<.05 FWE-corrected). Lower FA was also observed in those with TRS compared with UTRS in the superior longitudinal fasciculus (p<.05 FWE-corrected). However, post-hoc tests failed to survive corrections for multiple comparisons across the 12 post-hoc contrasts. No differences in MD, PD or RD were observed between groups. These data suggest that TRS is distinct from UTRS and that lower FA could act as a biomarker of treatment resistance in people with schizophrenia.

## Introduction

Pharmacological intervention is the most effective treatment for schizophrenia (Matheson et al., 2014). However, the rate of response to initial treatment with typical or atypical non-clozapine antipsychotics reaches only 60% to 80%, with response rates to a second non-clozapine antipsychotic as low as 16% (Agid et al., 2011; Elkis and Buckley, 2016). Resistance to first-line antipsychotic therapy accounts for some of the highest rates of hospitalisation and impaired functioning in mental health (Iasevoli et al., 2016; Lieberman and Murray, 2012) and is estimated to require USD 34 billion per annum in direct healthcare costs in the US alone (Kennedy et al., 2014). Furthermore, a recent meta-analysis (Siskind et al., 2017) revealed that clozapine, the most effective treatment for first-line therapy-resistant schizophrenia, is effective in only 40% of treatment-refractory individuals, suggesting that 12% to 20% of people with schizophrenia are ultra-treatment-resistant. Reflecting on these observations, researchers have posited that these three response levels represent three distinct subtypes of schizophrenia: treatment-responsive (first-line responders; FLR), treatment-resistant (clozapine responders; TRS) and ultra-treatment-resistant schizophrenia (clozapine-resistant; UTRS) (Farooq et al., 2013; Lee et al., 2015).

A growing body of research investigating potential differences in structural brain organisation between response subtypes of schizophrenia lends support to the concept of treatment-based categorical distinctions in this disorder and suggests that differences in white matter structure are present even at first episode of psychosis (Chen et al., 2018; Reis Marques et al., 2014). Reis Marques et al. (2014) reported that, at baseline, individuals with first-episode psychosis who failed to respond to a 12-week course of first-line antipsychotic therapy had lower fractional anisotropy (FA) than either healthy comparison subjects or responders in the left uncinate fasciculus, cingulum and superior longitudinal fasciculus and commissural tracts such as the corpus callosum (responders were indistinguishable from healthy comparison subjects). In support of these findings, Chen et al. (2018) identified white matter impairments in the right temporal and occipital lobes in resistant compared with responsive individuals with first-episode psychosis following one year of antipsychotic treatment. Differences in FA between responders and non-responders to first-line therapy also appear to persist throughout the course of disease. Widespread reductions in FA have been reported in patients who met criteria for TRS prior to commencing clozapine when compared to healthy subjects (Holleran et al., 2013). Researchers also identified lower FA in the body of the corpus callosum in those with clozapine-eligible schizophrenia compared with FLR (McNabb, Carolyn Beth et al., 2018), as well as reduced mean generalised FA within 12 white matter tracts in people with non-remitting compared with remitting schizophrenia (Huang et al., 2018). Two earlier cross-sectional studies of patients with chronic schizophrenia also reported that a poor response to antipsychotic treatment was associated with lower FA in the whole brain and in specific tracts such as the uncinate and superior longitudinal fasciculi (SLF) (Luck et al., 2011; Mitelman et al., 2006).

Although these studies provide mounting evidence for a role of white matter aberrations in treatment-resistant subtypes of schizophrenia, they have focused primarily on comparisons with healthy controls and differences between FLR and those with TRS (or poor treatment outcome), either omitting those with UTRS or absorbing them into the TRS subtype. If a diagnosis of schizophrenia really does divide into three distinct response subtypes, studies should be designed to distinguish between FLR, individuals with TRS and those with UTRS. Therefore, the current study sought to investigate white matter markers of treatment response in people with schizophrenia using the classification scheme proposed by Farooq et al. (2013) and Lee et al. (2015); specifically, categorising the disorder into FLR, TRS and UTRS subtypes.

## Experimental methods

### Participants

Details of participant recruitment have been described previously (Anderson et al., 2015; Goldstein et al., 2015; McNabb, C. B. et al., 2018). Individuals with a diagnosis of schizophrenia according to the Diagnostic and Statistical Manual of Mental Disorders (DSM-IV) were recruited from mental health services (a community mental health centre or forensic psychiatric inpatient unit) in Auckland, New Zealand. Participants were enrolled into one of three study arms (FLR, TRS or UTRS). Those who were responding well to first-line atypical antipsychotic monotherapy were assigned to the FLR group; response was defined as an improvement of positive symptoms according to standard practice and current treatment guidelines for schizophrenia (Lehman et al., 2004; McGorry, 2005). Those who had failed at least two previous six-to-eight-week trials of atypical antipsychotics and were receiving clozapine at the time of screening were assigned to the TRS group and participants who had failed at least two previous six-to-eight-week trials of atypical antipsychotics and had also failed an adequate trial of clozapine monotherapy (at least 8 weeks post titration (Mouaffak et al., 2006)) were assigned to the UTRS group. Patients in all treatment groups were required to be at most mildly ill, determined using the Clinical Global Impression (CGI) (which corresponds to a score of 3 out of 7 using this scale; 7 being the most extremely ill) to control for state effects (as otherwise differences in the TRS/UTRS groups could be ascribed to the presence of more severe symptoms). A healthy comparison group with no history of mental or neurological illness was recruited by advertising in the community. Age, sex and ethnicity were matched on a group basis. All participants were between the ages of 18 and 45 years. Exclusion criteria were as follows: history of traumatic brain injury (loss of consciousness for more than three minutes), neurological illness, significant physiological comorbidity or contraindication to magnetic resonance imaging (MRI). The study was approved by the Northern X Regional Ethics Committee (reference: NTX/09/05/042) and conducted in accordance with the Helsinki Declaration as revised 1989. All participants gave informed written consent - ability to give informed consent was judged by the treating psychiatrist.

Duration of psychosis and Positive and Negative Syndrome Scale (PANSS) scores (Kay et al., 1987) were assessed at study entry. Antipsychotic dose at the time of assessment was converted to chlorpromazine equivalents (CPZE) using formulae with power transformation (Andreasen et al., 2010). In the absence of a power formula for amisulpride, CPZE were calculated using expert consensus regarding antipsychotic dosing (Gardner et al., 2010). Participants also provided a urine sample, which was screened for the presence of amphetamine, methamphetamine, benzodiazepines, cocaine, opiates and tetrahydrocannabinol (Medix Pro-Split Integrated Cup, Multi Drug Screening Test; Sobercheck Ltd). Standardised premorbid intelligence quotient (IQ) scores were determined using a computerised version of the “spot the real word” task within the IntegNeuro test battery (Brain Resource Company, Sydney, Australia) (Baddeley et al., 1993; Gordon, 2003; Gordon et al., 2005). The sample size was based on previous studies examining the relationship between treatment outcomes and diffusion tensor imaging (DTI) measures in schizophrenia (Garver et al., 2008; Luck et al., 2011; Mitelman et al., 2009).

### Image acquisition and pre-processing

Scanning was performed using a 3T Siemens Magnetom Skyra (Siemens, Germany). A 32-channel head coil was used for the majority of scans; where the participant could not fit comfortably into this coil, a 20-channel head coil was used (n=5). T1-weighted images were acquired using a magnetization-prepared 180-degrees radio-frequency pulses and rapid gradient-echo (MPRAGE) sequence (Brant-Zawadzki et al., 1992). Acquisition parameters were as follows: repetition time (TR) 1900 ms; echo time (TE) 2.39 ms; inversion time (TI) 900 ms; flip angle 9°; repetition 1; acceleration factor 2; field of view (FOV) 230 mm; matrix 256 × 256; voxel size 0.9 × 0.9 × 0.8 mm. Diffusion-weighted images were acquired using an echo planar imaging (EPI) sequence with the following parameters: TR 8900 ms, TE 95 ms, FOV 240 mm, matrix 122 × 122, 2.0 mm slice thickness, isotropic voxel size 2.0 × 2.0 × 2.0 mm. An acceleration factor (GRAPPA) of 2 was used. Sixty-seven slices were acquired in the anterior to posterior direction with diffusion-weighting factor b=1000s/mm^2^ in 64 unique gradient-encoding directions. Eight scans without diffusion weighting (b=0) scans were also acquired. All scans were examined for artefacts including subject motion, excess signal-to-noise and incomplete acquisitions. No participants were excluded on this basis.

Measures of white matter integrity, including FA, mean diffusivity (MD), parallel (axial) diffusivity (PD) and radial diffusivity (RD) were obtained using Tract-Based Spatial Statistics (TBSS) (Smith et al., 2006), part of the FMRIB Software Library - FSL (Smith et al., 2004). FA images were created by fitting a tensor model to the raw diffusion data using FMRIBs Diffusion Toolbox (FDT), and then brain-extracted using the Brain Extraction Tool (BET) (Smith, 2002). All subjects’ FA data were then aligned to the FMRIB58_FA target image using the nonlinear registration tool FNIRT (Andersson et al., 2007) which uses a b-spline representation of the registration warp field (Rueckert et al., 1999) and transformed into Montreal Neurologic Institute (MNI)-152 standard space. Next, a mean FA skeleton representing the centres of all white matter tracts common to the group was created and thinned at a threshold of 0.3 to exclude low anisotropic regions. Each subject’s aligned FA data were then projected onto this skeleton. Anatomical localisation of significant white matter clusters was determined using the Juelich histological white matter labels and tractography atlases. The same approach was used for preprocessing of MD, PD and RD data.

### Statistical analysis

Whole-brain statistical analyses were performed using FSL’s Randomise tool. Threshold-free cluster enhancement was used for statistical comparisons, utilising a nonparametric permutation test in which group membership was permuted 5,000 times to generate a null distribution for each contrast. An analysis of variance (ANOVA) was employed to investigate whether any difference between groups existed in the data. Post hoc unpaired *t*-tests were then used to explore potential differences between the groups. Age and sex were demeaned prior to analysis and included as covariates of no interest in both ANOVA and post hoc analyses (Bose et al., 2009). A single regression correlation (Pearson) was also performed to investigate the association between white matter measures and daily antipsychotic dose in CPZE. All voxel-wise *p*-values were corrected for multiple comparisons using a Family-Wise Error (FWE) correction threshold of *p* = .05; post hoc tests were corrected for multiple comparisons using the False Discovery Rate (FDR). Both corrected and uncorrected post hoc *t*-test results are presented.

Analyses of socio-demographic and clinical characteristics were conducted using the Psych and DescTools packages for R (R Core Development Team, 2012; Revelle, 2017; Signorell et al., 2019). ANOVA was used to compare continuous variables with Bonferroni correction for multiple comparisons of post hoc tests. Fisher’s Exact test was used for comparisons of non-parametric data. FA images were created in FSLeyes and plots were created using the ggplot2 package in R (Wickham, 2016).

## Results

### Participants

A total of 70 participants (20 healthy controls, 18 FLR, 18 individuals with TRS and 14 with UTRS) were included in the final analysis. Demographic and clinical data are summarized in Table 1. There were no significant differences in gender, age, IQ or positive tests for THC between the groups. No differences in duration of illness, duration of untreated psychosis, PANSS total scores, or any of the subscales were observed between the groups with schizophrenia. Those with UTRS were receiving a higher dose of antipsychotics (measured in CPZE) at the time of scanning compared with both FLR and those with TRS; clusters demonstrating differences in FA between treatment groups were therefore correlated against CPZE to determine whether antipsychotic dose affected the results.

**Table 1.** Demographic data (mean (SD) unless otherwise stated) of study participants by treatment group.

|  | Controls<br>(n=20) | FLR<br>(n=18) | TRS<br>(n=18) | UTRS<br>(n=14) | Test Statistic |
| --- | --- | --- | --- | --- | --- |
| Sex (male/female) | 17/3 | 14/4 | 14/4 | 11/3 | *Fisher's Exact test (FET),*<br>*p = .933* |
| Age at scan (years) | 33.2 (8.4) | 32.2 (7.9) | 33.2 (8.2) | 34.6 (7.0) | $F(3,66) = .31, p = .818$ |
| Premorbid IQ (standardized scores) | -0.838 (1.286) | -0.781 (0.982) | -0.955 (1.170) | -0.885 (0.883) | $F(3,66) = .08, p = .971$ |
| Duration of illness (years) | - | 10.0 (7.9) | 12.5 (6.7) | 12.2 (4.8) | $F(2,47) = .73, p = .485$ |
| Duration of untreated psychosis (months) | - | 12.5 (14.7) | 11.1 (18.1) | 22.5 (27.5) | $F(2,47) = 1.46, p = .242$ |
| PANSS total score | - | 59.5 (11.0) | 59.6 (14.1) | 61.2 (10.9) | $F(2,47) = .09, p = .911$ |
| PANSS positive subscale score | - | 13.4 (5.36) | 11.9 (5.5) | 12.6 (5.1) | $F(2,47) = .36, p = .703$ |
| PANSS negative subscale score | - | 17.1 (5.75) | 18.8 (6.8) | 19.5 (7.6) | $F(2,47) = .57, p = .572$ |
| PANSS general subscale score | - | 28.9 (6.01) | 28.8 (6.2) | 29.1 (4.4) | $F(2,47) = .01, p = .993$ |
| Current prescribed antipsychotic | - | Olanzapine (n=8)<br>Risperidone (n=6)<br>Aripiprazole (n=3)<br>Amisulpride (n=1) | Clozapine (n=18) | Clozapine + amisulpride (n = 4)<br>Clozapine + aripiprazole (n = 4)<br>Clozapine + risperidone (n = 2)<br>Clozapine + quetiapine (n = 1)<br>Aripiprazole + quetiapine (n = 2)<br>Risperidone + quetiapine (n = 1) |  |
| Dose at time of scan (chlorpromazine equivalents) | - | 421.5 (191.6) | 438.2 (229.3) | 843.2 (377.4) | $F(2,47) = 12.03, p < .001$<br>**UTRS vs FLR, $p < .001$**<br>**UTRS vs TRS, $p < .001$** |
| Positive test for THC (n) | 0 | 3 | 1 | 3 | $FET, p = .096$ |

### TBSS results

An ANOVA between all study groups revealed a statistically significant effect for FA but not for MD, PD or RD. Pairwise comparisons between groups (FWE-corrected) revealed lower FA in those with TRS compared to healthy controls (figure 1) as well as those with UTRS (figure 2). Neither test survived corrections for multiple post hoc comparisons between groups using the FDR; however, the consistency of our findings with the literature to date suggests that this is most likely a result of small sample size as opposed to false positive results.

**Figure 1.**
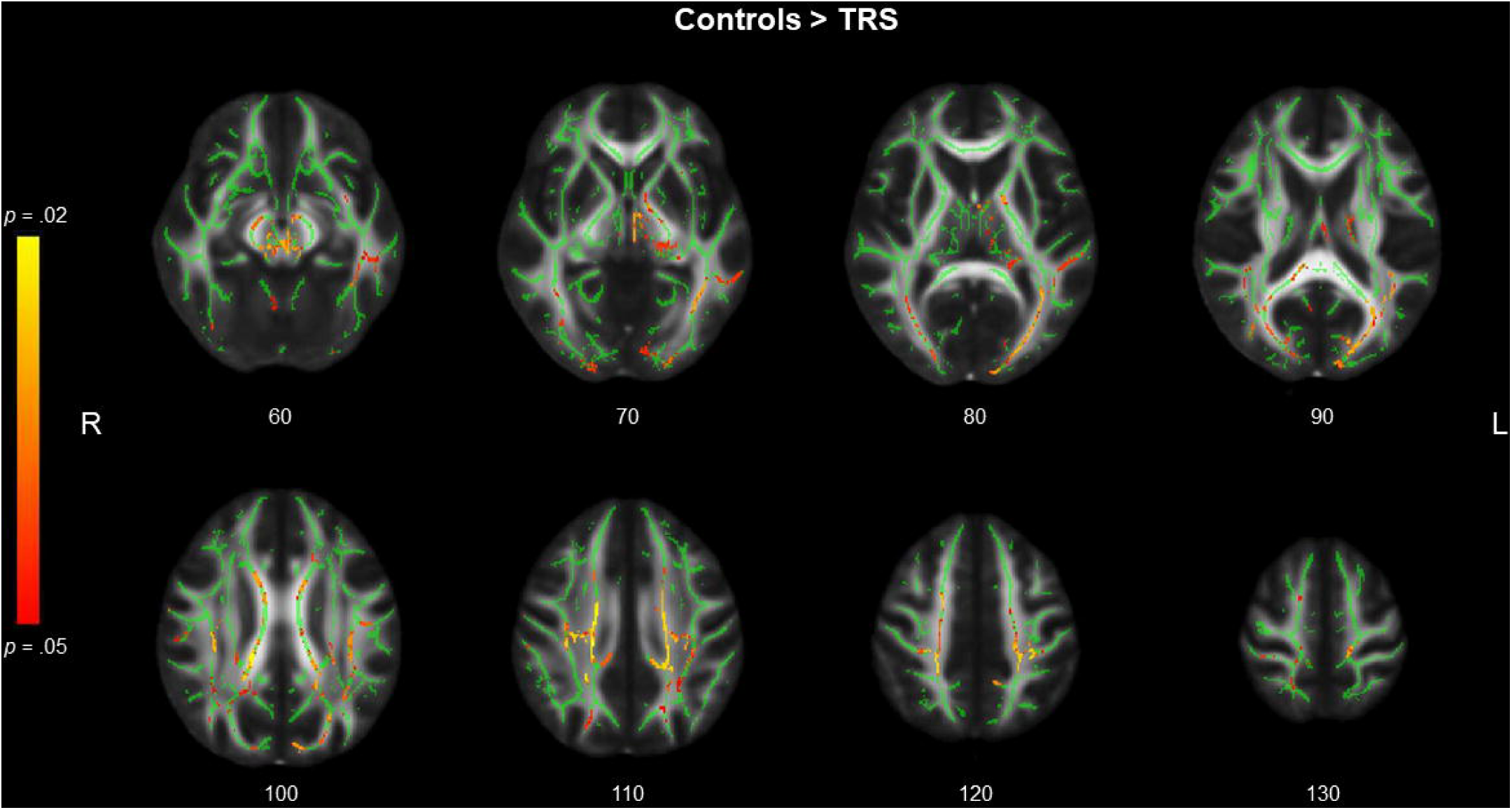
White matter maps showing lower fractional anisotropy (FA) in the treatment resistant schizophrenia (TRS) group compared to psychiatrically healthy controls (p < .05, FWE-corrected). Red-yellow voxels represent regions in which FA is significantly lower in the TRS group relative to healthy controls, overlaid on to the mean FA skeleton (green) and FMRIB58-FA image in standard MNI-152 space (radiologic view with Z coordinates). FWE = family-wise error; MNI = Montreal Neurological Institute.

**Figure 2.**
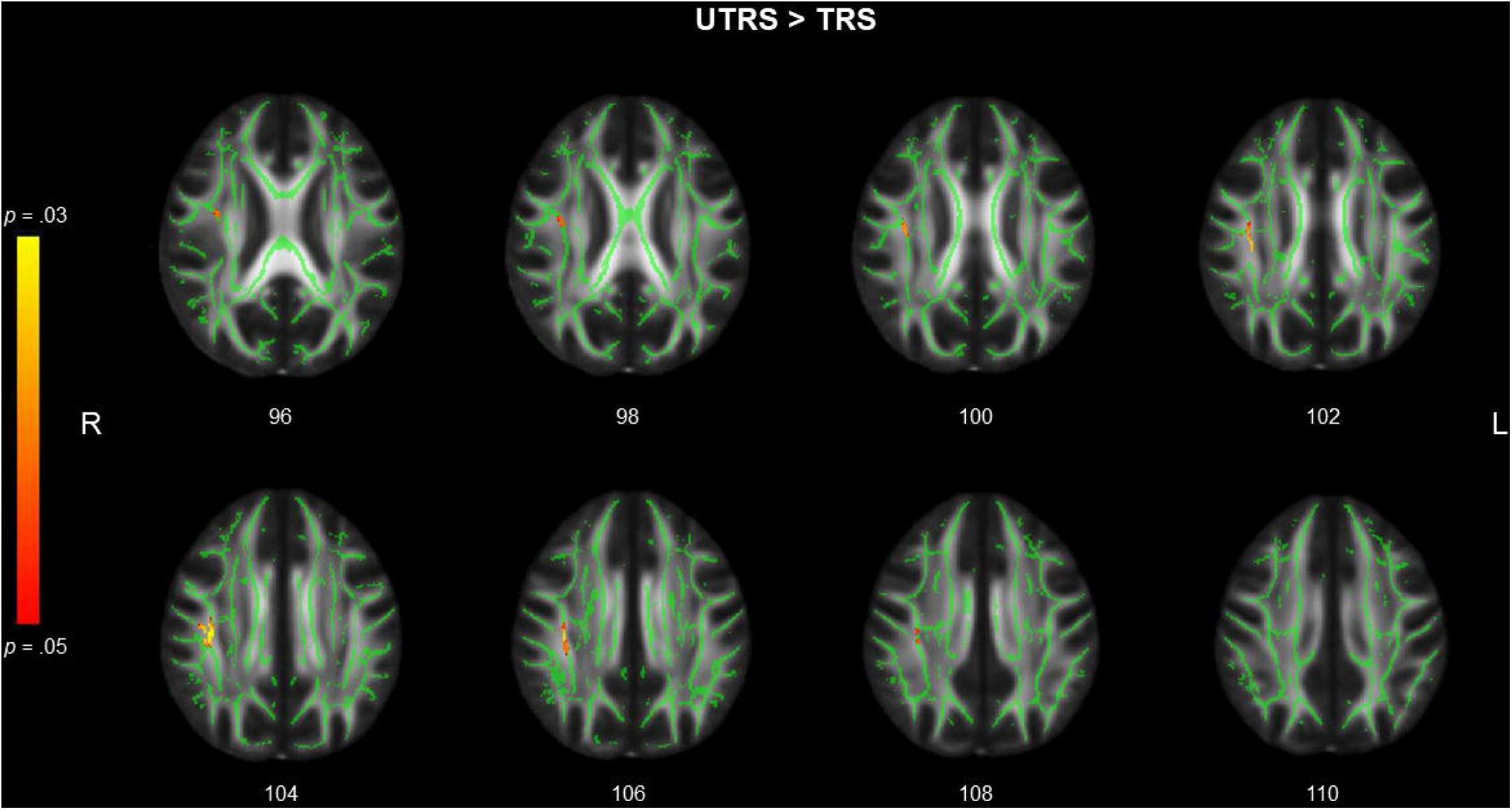
White matter maps showing lower fractional anisotropy (FA) in the treatment-resistant schizophrenia (TRS) group compared to the ultra-treatment-resistant schizophrenia (UTRS) group (p < .05, FWE-corrected). Red-yellow voxels represent regions in which FA is significantly lower in the TRS group relative to UTRS, overlaid on to the mean FA skeleton (green) and FMRIB58-FA image in standard MNI-152 space (radiologic view with Z coordinates). FWE = family-wise error; MNI = Montreal Neurological Institute.

Clusters with lower FA in those with TRS are shown in figure 3 and described further in table 2. Those with TRS had lower FA than controls in the superior longitudinal fasciculus (bilateral), corpus callosum, thalamic radiation (bilateral), corticospinal tract (bilateral), internal capsule (left), corona radiata (bilateral) and fronto-occipital fasciculus (bilateral), among other regions. In contrast, differences between those with TRS and UTRS were confined to the right superior longitudinal fasciculus (including the temporal component of the superior longitudinal fasciculus).

**Table 2.** Description of white matter clusters showing lower fractional anisotropy (FA) in those with treatment-resistant schizophrenia (TRS) compared with either psychiatrically healthy controls or those with ultra-treatment-resistant schizophrenia (UTRS). Regions are shown for the Juelich histological white matter labels and tractography atlases and are ordered according to greatest probability of the cluster residing in each region (averaged across all voxels in the cluster). Regions with probability >1% only are presented here.

| Cluster Index | White matter tract/region |  | Cluster size<br>(number voxels) | Peak *p*<br>value | Peak voxel (MNI-152<br>coordinates, mm) |  |  |
| --- | --- | --- | --- | --- | --- | --- | --- |
|  | JHU ICBM-DTI-81 White-Matter Labels | JHU White-Matter Tractography Atlas |  |  | x | y | z |
| **UTRS>TRS** |  |  |  |  |  |  |  |
| **Cluster:**<br>**1** | Superior longitudinal fasciculus (R) | Superior longitudinal fasciculus<br>(temporal part, R)<br>Superior longitudinal fasciculus (R) | 395 | 0.97 | 50 | 104 | 103 |
| **CON>TRS** |  |  |  |  |  |  |  |
| **Cluster:**<br>**7** | Body of corpus callosum<br>Superior longitudinal fasciculus (R)<br>Splenium of corpus callosum<br>Superior corona radiata (R)<br>Posterior thalamic radiation (including optic<br>radiation, R)<br>Posterior corona radiata (R) | Superior longitudinal fasciculus (R)<br>Inferior fronto-occipital fasciculus (R)<br>Forceps major<br>Inferior longitudinal fasciculus (R)<br>Corticospinal tract (R)<br>Superior longitudinal fasciculus<br>(temporal part, R) | 6150 | .022 | 74 | 92 | 100 |
| **6** | Posterior thalamic radiation (include optic<br>radiation) (L)<br>Superior longitudinal fasciculus (L) | Inferior longitudinal fasciculus (L)<br>Inferior fronto-occipital fasciculus (L)<br>Forceps major | 4467 | .029 | 124 | 67 | 75 |
|  | Sagittal stratum (include inferior longitudinal fasciculus and inferior fronto-occipital fasciculus, L) | Superior longitudinal fasciculus (L)<br>Superior longitudinal fasciculus (temporal part, L) |  |  |  |  |  |
| **5** | Cerebral peduncle (bilateral)<br>Posterior limb of internal capsule (L)<br>Anterior limb of internal capsule (L)<br>Superior cerebellar peduncle (R)<br>Fornix (cres) / Stria terminalis (L)<br>Inferior cerebellar peduncle (R)<br>Superior cerebellar peduncle (L) | Anterior thalamic radiation (L)<br>Corticospinal tract (bilateral)<br>Anterior thalamic radiation (R) | 3747 | .028 | 88 | 96 | 57 |
| **4** | Superior longitudinal fasciculus (L)<br>Body of corpus callosum<br>Superior corona radiata (L)<br>Splenium of corpus callosum<br>Posterior corona radiata (L)<br>Anterior corona radiata (L) | Superior longitudinal fasciculus (L)<br>Superior longitudinal fasciculus (temporal part, L) | 3124 | .025 | 110 | 96 | 110 |
| **3** |  | Inferior longitudinal fasciculus (L) | 120 | .049 | 102 | 52 | 113 |
| **2** | Superior longitudinal fasciculus (R) | Superior longitudinal fasciculus (R)<br>Superior longitudinal fasciculus (temporal part, R) | 116 | .049 | 50 | 79 | 90 |
| **1** | External capsule (L) | Inferior fronto-occipital fasciculus (L)<br>Uncinate fasciculus (L) | 5 | .050 | 118 | 142 | 70 |

**Figure 3.**
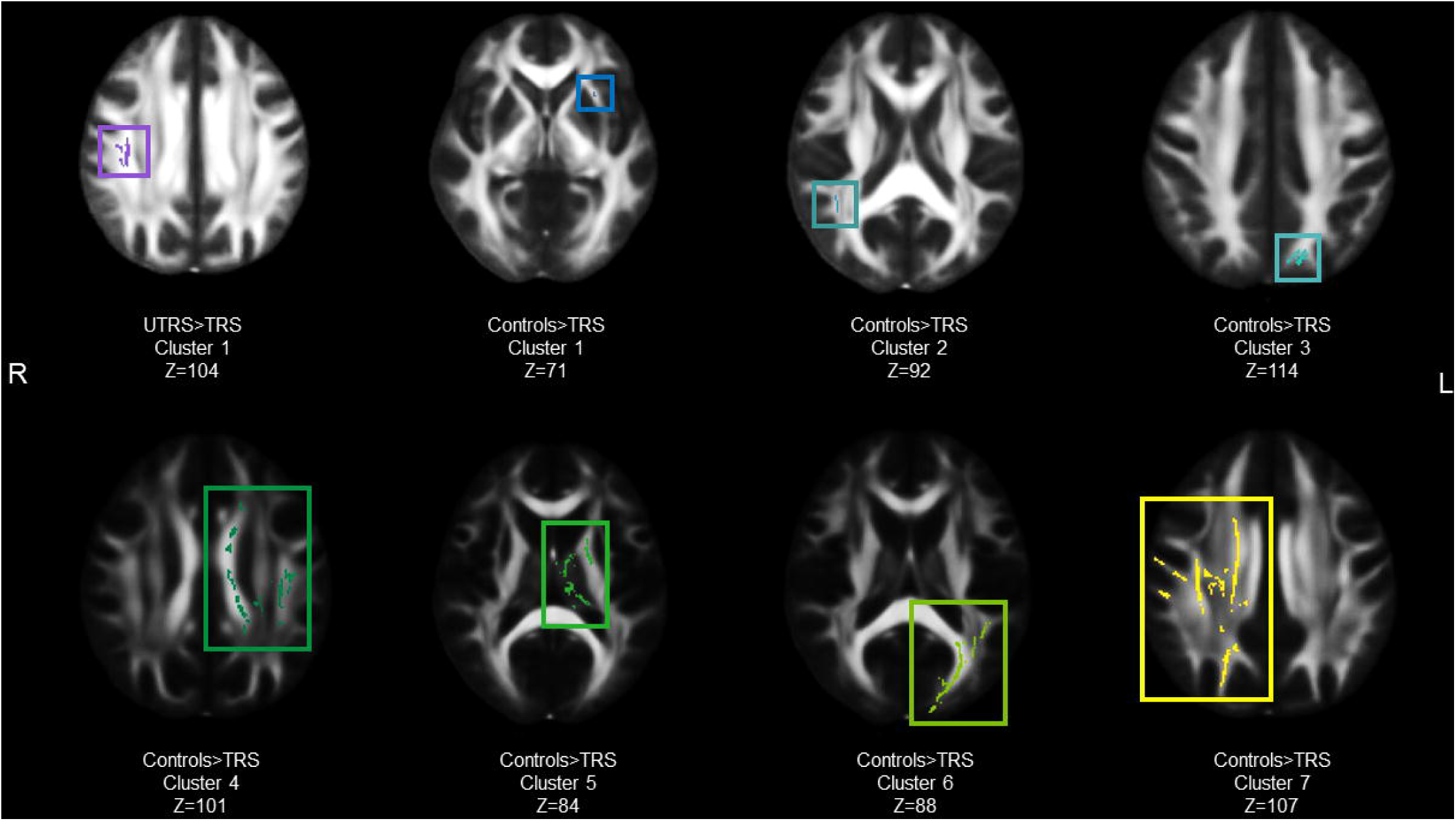
Voxel clusters in which individuals with treatment-resistant schizophrenia (TRS) exhibited lower fractional anisotropy (FA) than psychiatrically healthy controls or those with ultra-treatment-resistant schizophrenia (UTRS).

To further interrogate the differences between groups, mean FA for each cluster was determined for every participant (figure 4). FA was lowest in those with TRS for all clusters and, for the most part, was similar for FLR and those with UTRS, both of which had slightly lower FA than healthy controls. The exception was for the single cluster exhibiting lower FA in those with TRS compared with UTRS, in which the effect appears to be driven by a combination of low FA in those with TRS and high FA in those with UTRS.

**Figure 4.**
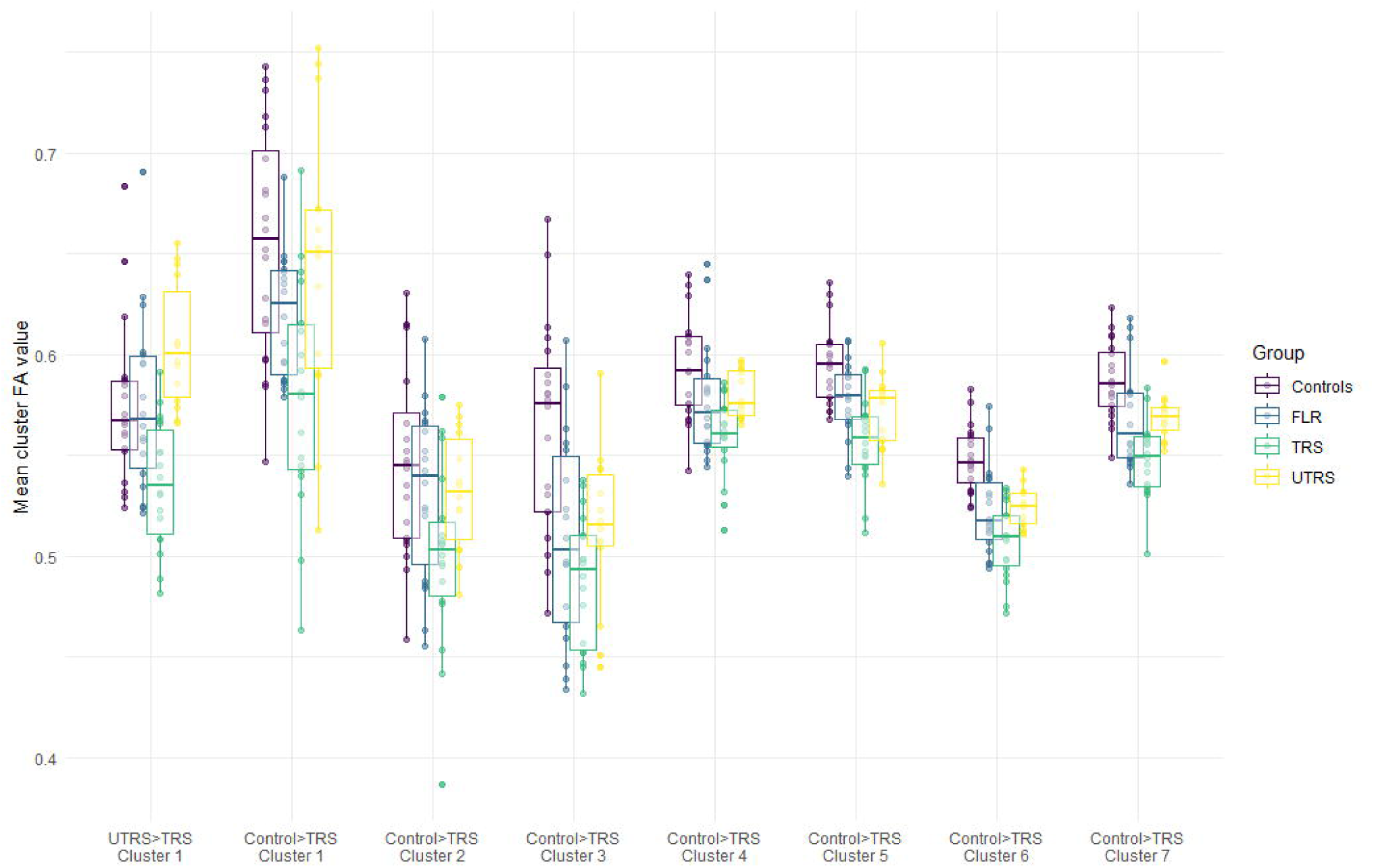
Mean fractional anisotropy (FA) for clusters in which the group with treatment resistant schizophrenia (TRS) had significantly lower FA than healthy comparison subjects or those with ultra-treatment-resistant schizophrenia (UTRS). Individual subjects’ mean FA values are represented by points, with group data represented by each box - lines through each box represent the group median; the upper and lower “hinges” correspond to the first and third quartiles (the 25th and 75th percentiles). Data beyond the end of the whiskers are outliers (as specified by Tukey).

### Correlation with clinical characteristics

As CPZE demonstrated differences between treatment groups, we ran additional regression analyses investigating the relationships between CPZE and white matter measures. There was no area identified where FA, MD, PD or RD correlated with antipsychotic daily dose in CPZEs. Likewise, no statistically significant relationship between cluster FA and antipsychotic dose measured in CPZE was identified when measured independently for each cluster (figure 5). Similar analyses were conducted for PANSS total score and age at onset of psychosis in those with schizophrenia and no significant relationships to cluster FA values were identified (see supplementary figures S1 and S2, respectively).

**Figure 5.**
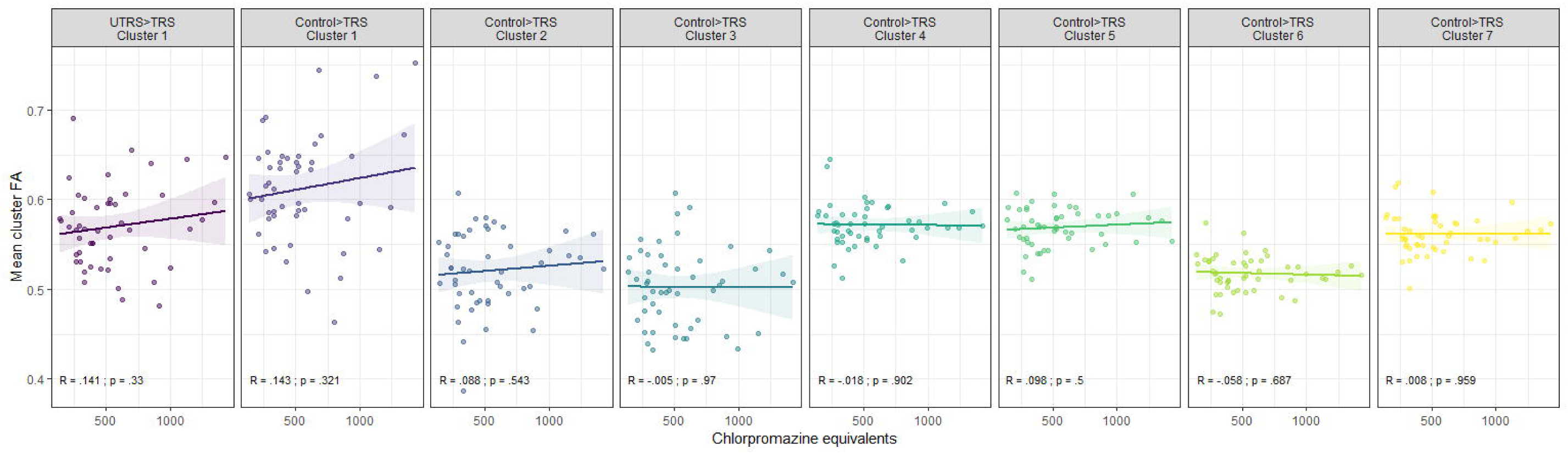
Relationship between mean cluster fractional anisotropy (FA) and antipsychotic dose measured in chlorpromazine equivalents (CPZE) in those with schizophrenia. Pearson’s correlation (R) and uncorrected *p* values are presented for each cluster.

### Correlation with socio-demographic characteristics

As previous work found significant associations between IQ and FA (Luders et al., 2007; Luders et al., 2011; Navas-Sánchez et al., 2014), for each cluster, we ran correlations between mean FA and standardised (premorbid) IQ. IQ correlated with mean FA in two clusters (figure 6), although only one survived corrections for multiple comparisons across clusters (Bonferroni-corrected *p* threshold .007). The significant correlation was for a cluster spanning only 5 voxels (located in the left external capsule / inferior fronto-occipital fasciculus / uncinate fasciculus) whereby those with TRS had lower FA than healthy controls. The remaining clusters showed no relationship between FA and IQ.

**Figure 6.**
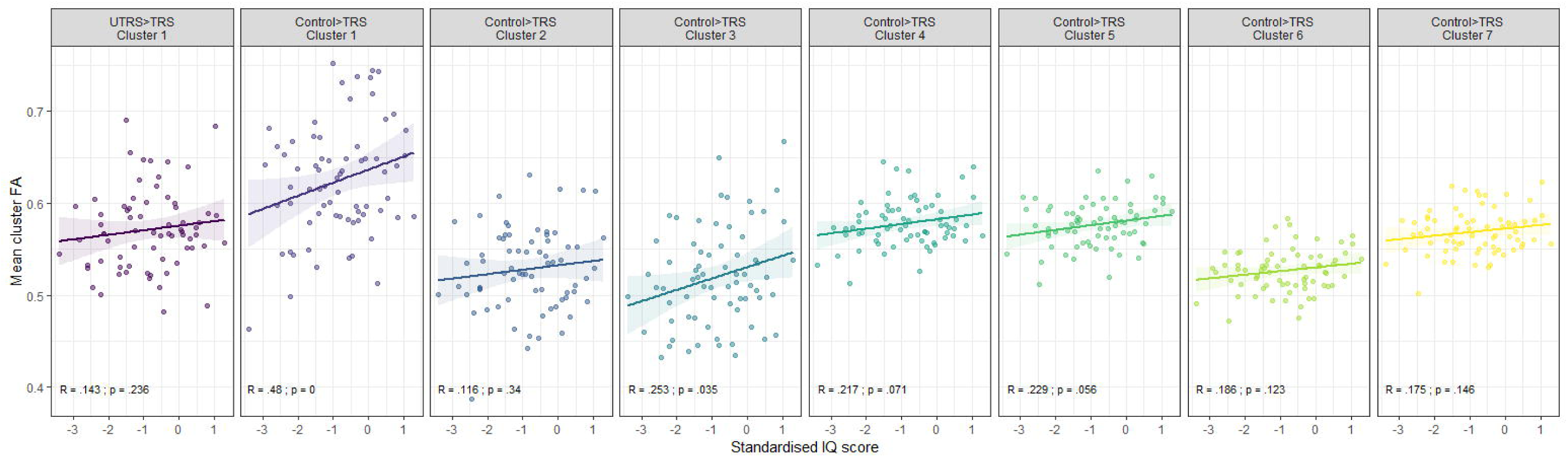
Relationship between mean cluster fractional anisotropy (FA) and standardised (premorbid) IQ. Pearson’s correlation (R) and uncorrected *p* values are presented for each cluster.

## Discussion

Results from studies investigating white matter integrity in people with schizophrenia have increasingly contributed toward a consensus that people who fail to respond to first-line antipsychotic therapy exhibit greater disruptions in FA than treatment responders (Chen et al., 2018; Huang et al., 2018; McNabb, Carolyn Beth et al., 2018; Reis Marques et al., 2014). However, schizophrenia encompasses two distinct subtypes of treatment resistance – clozapine responsive treatment resistance (TRS) and clozapine resistant treatment resistance (UTRS). This was the first study to explicitly define and compare these two subtypes in addition to FLR and healthy controls to investigate possible white matter markers of treatment resistance in people with schizophrenia. Our results suggest that TRS and UTRS are distinct from one another in their white matter profiles, TRS exhibiting lower FA than both healthy controls and UTRS. Therefore, resistance to the clinical benefits of first-line antipsychotic drugs and subsequent sensitivity to clozapine may be attributable to global disruptions in FA. By comparison, those with UTRS had FA comparable to FLR and healthy controls, suggesting that white matter disruptions play a less pivotal role in treatment resistance for these individuals.

Prominent regions of white matter disruptions in our TRS cohort included the superior longitudinal fasciculus (including the temporal region of this tract), body and splenium of the corpus callosum and forceps major, corticospinal tract, superior and posterior corona radiata, posterior limb of the internal capsule, anterior and posterior thalamic radiation, the external capsule, uncinate fasciculus, inferior fronto-occipital fasciculus and cerebral peduncle. Our findings of widespread white matter aberrations in those with TRS are in agreement with previous studies investigating white matter integrity in responders and non-responders to first-line therapy. Luck et al. (2011) identified reduced FA in people with chronic schizophrenia experiencing poor response compared with healthy controls within the superior longitudinal fasciculus and uncinate fasciculus but not the cingulum, with FA reductions more pronounced in the poor outcome compared with good outcome group (Luck et al., 2011). Likewise, Reis Marques et al. (2014) found that those who went on to develop a poor response to antipsychotic treatment exhibited decreased FA across multiple white matter regions, including the left uncinate, cingulum and superior longitudinal fasciculus and corpus callosum, compared to healthy controls. The corpus callosum is the fundamental white matter structure conferring interhemispheric connectivity. Lower FA in the forceps major, splenium and body of the corpus callosum in our TRS group is in agreement with previous studies investigating treatment resistance, as well as schizophrenia more generally. Decreased FA in the corpus callosum is consistently reported in patients with schizophrenia (Bora et al., 2011; Foong et al., 2000; Kong et al., 2011; Kubicki et al., 2008; Patel et al., 2011) and more specifically chronic schizophrenia (Douaud et al., 2007; Koch et al., 2010; Kubicki et al., 2008; Mitelman et al., 2009; Miyata et al., 2010; Rotarska-Jagiela et al., 2008). Mitelman et al. observed lower average FA in the corpus callosum of individuals with schizophrenia with poor-outcome compared with good outcome (Mitelman et al., 2009). Lower FA in the body of the corpus callosum has also been observed in people with schizophrenia who are eligible for clozapine compared with those responding well to first-line antipsychotic therapy (McNabb, Carolyn Beth et al., 2018). Though we did not detect differences between patient groups in the current study, FA was lowest in the group with TRS throughout the corpus callosum body, splenium and forceps major, suggesting that callosal aberrations are most severe in this population.

Our finding of lower FA in the fronto-occipital fasciculus may be driven by disruptions in myelination in those with TRS. A recent study investigating magnetic resonance markers of myelination found lower myelin water fraction in people with schizophrenia compared with healthy controls in this region, although no difference between resistant and responsive patients were observed (Vanes et al., 2018). Myelin water fraction within the corpus callosum was also associated with cognitive control in those with schizophrenia. In their study, Vanes et al. (2018) explicitly excluded individuals receiving clozapine to avoid the introduction of subgroups including ultra-treatment-resistant schizophrenia. However, we have demonstrated that this subgroup exhibits white matter patterns quite distinct from TRS, suggesting that distinguishing TRS from UTRS provides useful information about the treatment response patterns of individuals with schizophrenia.

Although we did not demonstrate significant differences in white matter between all schizophrenia cohorts and healthy controls in the current analysis, a previous analysis of the same participants identified lower white matter volume in all schizophrenia groups compared with controls (Anderson et al., 2015). Therefore, the lower (but not statistically significant) FA observed in FLR (and to a lesser extent those with UTRS) compared with healthy controls could indicate a less severe but still clinically relevant reduction in FA in these individuals.

All participants in the TRS group and the majority of participants in the UTRS groups were receiving clozapine at the time of scanning. Previous studies have associated both a reduction in white matter volume (Molina et al., 2005) and increase in FA (Ozcelik-Eroglu et al., 2014) with the use of clozapine. As our results demonstrate lower FA in those on clozapine monotherapy and similar levels of FA in those with UTRS (receiving augmented clozapine therapy) and FLR, it is unlikely that our effects are driven by exposure to clozapine. However, if FA is lowest in those with TRS and clozapine increases FA, this could be a mechanism by which clozapine improves symptoms of the disorder.

Antipsychotic dose is also unlikely to have driven the effects observed in this study. As the UTRS group were receiving significantly higher doses of antipsychotics (measured in CPZE) compared with both FLR and those with TRS, we ran correlations within the whole brain and significant FA clusters to determine the effects of CPZE. No significant effects were observed in either analysis, suggesting that the lower FA in those with TRS is independent of antipsychotic exposure.

We did, however, identify a relationship between IQ and mean FA in one of the seven clusters (located in the external capsule) demonstrating differences between TRS and healthy controls. IQ has been shown to correlate with white matter integrity in the external capsule in previous research (Navas-Sánchez et al., 2014); however, as the cluster demonstrating a significant effect contained only 5 voxels, it is unlikely that IQ is driving the white matter differences between those with TRS and healthy controls. In addition, the remaining clusters showed no such relationship, therefore we suggest that the relationship between IQ, treatment resistance and measures of white matter integrity is weak at best.

This study benefits from a well characterised cohort of participants, well matched for clinical and demographic variables including gender, age, duration of illness, duration of untreated psychosis and premorbid IQ. Participants taking clozapine had well-established TRS and patient groups were stabilized on their current treatment. Inclusion of patients with mild symptoms provides us with confidence that the differences in white matter integrity observed in those with TRS are unlikely to be the result of more severe active symptoms in this cohort. Correlation analysis with total PANSS scores supports this theory.

The study’s cross–sectional design imparts a degree of uncertainty about the underlying cause of observed differences. However, these findings are in line with previous work highlighting differences between controls and non-responders to antipsychotic therapy, suggesting the effects are robust. Therefore, although our post hoc analyses did not survive corrections for multiple comparisons between groups, their concordance with previous work provides a strong foundation on which a theory attributing treatment resistance to white matter abnormalities can be built.

In summary, these results suggest that microstructural white matter abnormalities in people with schizophrenia are more pronounced in those with TRS than those who respond well to first-line antipsychotics and those with UTRS and that these aberrations exist despite effective treatment with clozapine. Most critically, we have demonstrated that TRS and UTRS represent two distinct subtypes of treatment-resistant schizophrenia. As differences between TRS and UTRS were driven by low FA in those with TRS combined with high FA (compared to healthy controls) in those with UTRS, and previous work in this population found disruptions in functional connectivity in this cohort, ultra-treatment resistance is likely driven by different underlying mechanisms to TRS. We hope these findings, in combination with the current literature, will encourage researchers to delineate between treatment-resistant and ultra-treatment resistance schizophrenia in the future.

## Data Availability

This data is confidential and cannot be shared

